# Exploring Sleep, Inflammation and Myocardial Infarction Recurrence in Aging

**DOI:** 10.64898/2026.01.14.26343929

**Authors:** Sophie Hirsch, Christopher M. Smith, Carolyn E. Horne

## Abstract

**Background:** Despite cardiovascular disease being a leading cause of death globally, secondary prevention strategies following myocardial infarction are under-researched compared to primary prevention.

**Objectives:** To explore relationships between indicators of sleep quality, inflammation, and myocardial infarction recurrence and the conditional role of psychological stress in these pathways to assess evidence for reducing MI recurrence by improving sleep quality and implementing stress management techniques.

**Methods:** A secondary analysis (*N* = 156) of cross-sectional study data was conducted. Participants’ mean age was 65 years and all had experienced one or more myocardial infarctions within the previous 3-7 years. Sleep quality was measured using the Pittsburgh Sleep Quality Index. Inflammatory markers were collected via blood assay and included C-reactive protein, interleukin-6, and tumor necrosis factor-alpha. Psychological stress was evaluated using a self-reported Likert-scale question. Correlational analysis and conditional path analyses examined relationships between variables of interest.

**Results:** Significant associations between sleep quality, inflammation, psychological stress, and MI recurrence were observed. Sleep quality predicted elevated inflammation markers. Psychological stress moderated the relationship between sleep quality and MI recurrence, with higher stress amplifying the risk. Sleep quality did not directly predict MI recurrence.

**Conclusion:** Findings suggest that improving sleep quality may be an effective secondary prevention strategy for reducing MI recurrence. The effect may be increased when combined with targeted stress management interventions. Further research is warranted to explore biobehavioral mechanisms underlying these associations to develop targeted interventions for aging individuals at risk.

## Introduction

Cardiovascular disease remains a leading cause of death globally, with older adults being particularly vulnerable to its effects.^1^ As individuals age, maintaining cardiovascular health becomes increasingly important due to increasing risk factors including myocardial infarction (MI) or its recurrence.^1–3^ However, secondary prevention strategies, particularly those embracing holistic health principles, remain largely underexplored. This study addressed the gap by investigating the complex relationships between sleep quality, perceived stress, and markers of inflammation in an aging population who experienced MI recurrence. Inflammatory markers such as Interleukin-1 beta (IL-1β) and C-reactive protein (CRP) are well-documented predictors of recurrent MI. Furthermore, sleep disturbances may play a role in exacerbating the chronic inflammatory processes that occur in aging.^4,5^ Thus, this research is situated within a framework of holistic aging, which emphasizes health and well-being to address complex challenges in aging.^6^

Key factors linked to cardiovascular health include smoking cessation, increased physical activity, improved nutrition, weight management, cholesterol control, blood pressure regulation, diabetes management, and sleep quality.^7^ Sleep encompasses multiple dimensions relevant to cardiovascular health, including duration, timing, regularity, and quality. The present study focuses specifically on sleep quality as one modifiable component within this broader sleep–cardiovascular framework.^8^ These behaviors and clinical targets support health and quality of life in aging individuals.^9,10^ Beyond modifying these risk factors, the role of inflammation is significant in cardiovascular related prognosis, often transitioning to chronic inflammatory states as one ages.^5^

Chronic inflammation is a major risk factor for cardiovascular disease and can be more pronounced in aging individuals due to longstanding and continuous systemic inflammation.^2^ This process is referred to as *inflammaging*, a term coined by Franceschi et al.^5^ One mechanism underlying inflammaging is chronic stimulation of the immune system. For example, following an MI, proinflammatory cytokines, such as interleukin-6 (IL-6) and tumor necrosis factor-alpha (TNFα) are released.^11^ These cytokines are involved in the development and progression of atherosclerosis, which may contribute to recurrent MIs. The intrinsic stress that occurs during an MI excites cytokines, including IL-1β, IL-6, CRP, and TNFα, producing an immune response,^1,5^ with levels increasing measurably within 24 hours.^11^ The presence of these cytokines predicts subsequent MI occurrences.^12^ In other words, once an individual has experienced their first MI, elevated levels of the same cytokines increase the risk of future MI, especially in older individuals.^2,5^ Therefore, exploring strategies to reduce levels of specific cytokines is crucial for supporting individuals who have experienced an MI and are older.

One of the essential secondary strategies for improving and maintaining cardiovascular health is targeting sleep quality.^1,7^ Unfavorable sleeping outcomes, including poor sleep quality and inadequate sleep duration are associated with numerous physiological and psychological health risks, particularly, cardiovascular disease.^13,14^ Martin et al.^1^ reported that older adults were less likely to report shorter sleep time (< 7 hours). However, Huang et al.^15^ found that sleep irregularity doubled the risk of heart disease over 5 years in participants (age, *M* = 69 years). These data suggest that in addition to the number of hours slept, sleep quality is a meaningful factor in aging populations. Beyond sleep duration and regularity, growing evidence indicates that poor subjective sleep quality, including fragmented sleep and reduced sleep efficiency, is independently associated with adverse cardiovascular outcomes in older adults and populations at elevated cardiovascular risk.^8,16^ While sleep quality is defined by both subjective satisfaction and objective measures and varies, sleep quality typically becomes less optimal with an overall reduction in quality as we age.^17,18^ Nelson et al.^19^ defined sleep quality as individuals’ self-satisfaction with aspects of the sleep experience consisting of self-efficiency, sleep latency, wakefulness after sleep onset and sleep architecture. Each component measures a different aspect of sleep quality and is defined or measured differently.

Factors influencing sleep quality include physiological variables (i.e., age, body mass index, existing health conditions), psychological variables (i.e., depression, anxiety, stress), and environmental conditions (i.e., electronic device usage, social interactions, ambient room temperature), that have been increasingly linked to inflammatory processes.^2,5,20^ In addition to psychological stress, physiological stress, such as chronic pain or effects from medications, may impact overall sleep quality.^21^ Medication effects represent a related but distinct consideration in this population, as many commonly prescribed cardiovascular and non-cardiovascular medications can influence sleep onset, maintenance, or daytime alertness, thereby contributing to overall sleep quality among older adults (e.g., beta-blockers, antidepressants, and other agents linked with fatigue or sleep disturbances).^22^

Sleep quality is closely associated with chronic inflammation, a significant contributor of cardiovascular disease.^15,16,23^ Disturbances in sleep are often accompanied by elevated levels of pro-inflammatory cytokines and a reduction in anti-inflammatory cytokines, leading to an imbalance that can promote chronic inflammation and elevated cardiovascular risk in aging.^2^ A meta-analysis examining the relationship between insomnia and the incidence of MI found that individuals who slept for only 5 hours had a 1.69-fold increased risk of experiencing an MI compared to those who slept 7-8 hours.^13^ Additionally, difficulties in both initiating and maintaining sleep were associated with a 1.13-fold increase in relative risk of MI. The meta-analysis concluded that there is a significant association between insomnia and increased risk of MI, with a 2.06 increase in relative risk in individuals > 65 years old.

The relationship between sleep quality and inflammation contributes to MI recurrence and is bi-directional.^24,25^ Chronic inflammation has the potential to disrupt sleep patterns, leading to sleep disturbances. In turn, sleep disturbances can exacerbate inflammation. This cyclical relationship may amplify the risk of MI recurrence. Irwin^24^ proposed a model in which sleep disturbances (infrequent sleep, too short or too long sleep, or poor sleep quality) activate the release of adrenocorticotropic hormone and cortisol into systemic circulation via the pituitary gland and the hypothalamus. This process signals leukocytes to produce proinflammatory cytokines such as IL-1β, IL-6, and TNFα while decreasing circulating anti-inflammatory cytokines such as interferon A and B. Thus, this suggests that sleep disturbances can disrupt hormonal balance and immune function, making individuals more susceptible to illness due to temporarily weakened immune function. Furthermore, if cytokines are released through the hypothalamic pituitary adrenal (HPA) axis due to diminished sleep quality, chronic sleep disturbances may lead to sustained HPA activation and increased release of inflammatory markers, potentially resulting in MI recurrence.

In this study, we investigated relationships between CRP, IL-1β, IL-6, TNFα, sleep quality, and MI recurrence. CRP is an acute pro-inflammatory protein that can increase up to 1,000-fold in an inflammatory setting.^26^ High levels of CRP are linked to sleep deprivation, excessive sleep duration, and poor sleep quality.^4,27^ Elevated CRP levels also strongly predict cardiovascular disease and low-density lipoproteins.^28^ TNFα is a “master regulator” pro-inflammatory cytokine involved in regulating inflammation in conditions such as rheumatoid arthritis,^29^ Crohn’s disease,^30^ multiple sclerosis,^31^ and uveitis.^32^

Current research underscores the strong associations between sleep behaviors and inflammation. TNFα levels are significantly elevated following sleep-deprivation^33^ and anti-TNFα therapy may temporarily improve self-reported sleep quality.^34^ IL-1β, an anti-inflammatory cytokine, is non-specific to antigen recognition^35^ and increases following sleep deprivation.^36^ Similarly, IL-6 is a nonspecific inflammatory marker that acts as a pro and anti-inflammatory cytokine in response to infection.^37^ Like TNFα, CRP, and IL-1β, IL-6 levels rise with sleep-deprivation and contribute to HPA activation, including the release of inflammatory and a decrease in anti-inflammatory inducible immunity-related GTPases proteins.^25^ However, these return to normal following a short nap.^38^ IL-6 levels are higher when sleep quality is poor.^39^

Psychological factors such as stress also play a role in sleep quality and holistic aging. ^17,18^ This is compounded by the psychological stressors that impact aging individuals such as depression, grief, and loneliness. Previous studies have confirmed that psychological stress or perceived stress directly affect the HPA axis.^40,41^ Similar to effects on sleep, dysregulation of the HPA axis signals cytokine release. Chronic stress can also dysregulate the HPA axis, leading to sustained cortisol release.^40,41^ Over time, this prolonged stress response promotes inflammation, worsening health outcomes. Daily stressors can lead to sleep problems and establish a feedback loop where stress causes poor sleep and in turn exacerbates stress and inflammation.^42^

The interplay between stress and inflammation may buffer certain health outcomes. For example, Tomfohr and colleagues^43^ found that CRP and TNFα concentrations were elevated in patients with cardiovascular disease who reported poor sleep but only if they also experienced social stress. Poor sleep quality and increased levels of psychological stress are associated in aging individuals with cardiovascular disease^44,45^ as well as stress and inflammation^4,46^ and inflammation and sleep.^24^ The complex relationship between these variables and their impact on health outcomes are largely unexplored.

This study was guided by Irwin’s model^24^ of sleep and immune function. We incorporated MI recurrence as an outcome and condition. We aimed to explore the relationships between sleep quality, inflammation, and MI recurrence and the conditional role of psychological stress in these pathways. Specifically, we examined whether sleep quality predicted MI recurrence through inflammation and tested the interaction effect of stress on the relationship between sleep quality and inflammation. The predictive model is shown in Figure 1.

**Figure 1.**
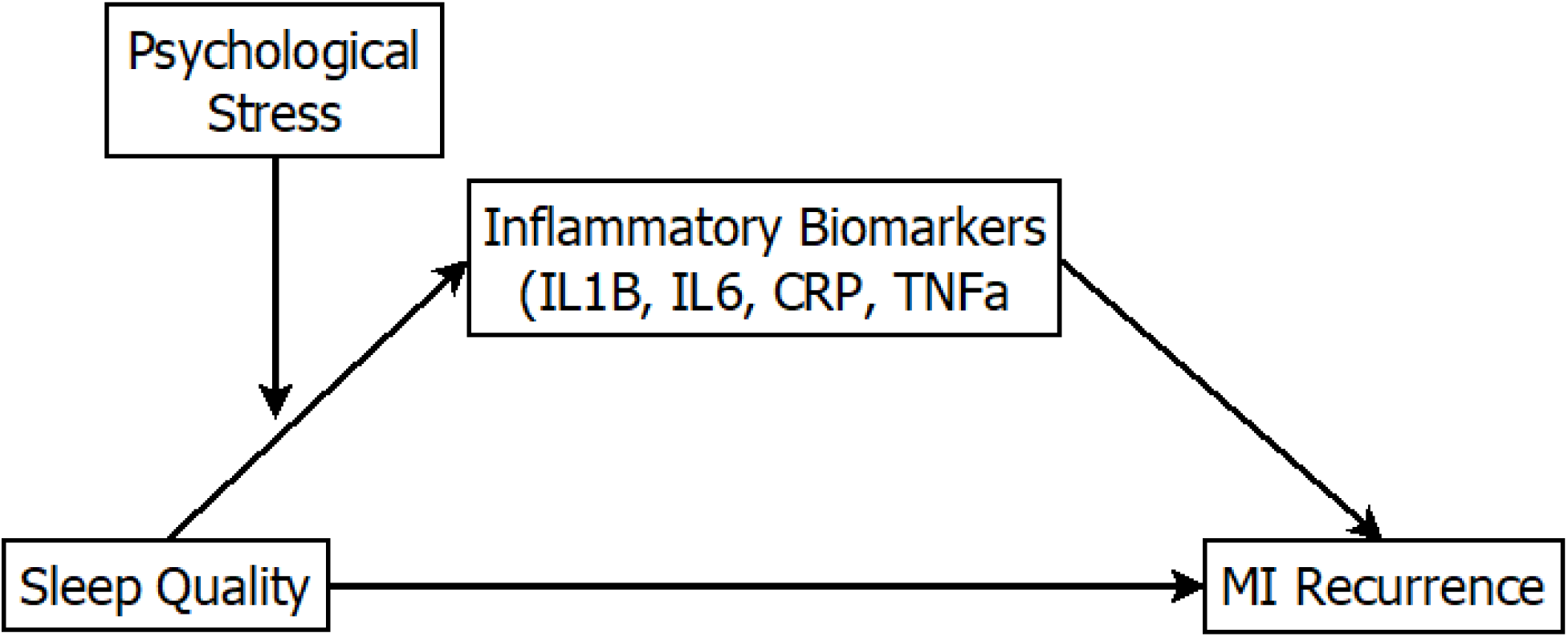
Proposed Model: Sleep is hypothesized to predict MI recurrence directly and indirectly via inflammatory biomarkers (IL-1ß, IL-6, CRP and TNFα). The relationship between sleep and inflammatory biomarkers is moderated by stress. *Note.* MI = Myocardial infarction.

## Methods

### Design

This study was a secondary analysis of a published cross-sectional study.^47^ Recruitment procedures and inclusion criteria are described briefly below. The study was approved by the Institutional Review Board.

### Participants

The parent study was comprised of *N =* 156 adults who had experienced at least one MI within the previous 3-7 years. Site collaborators at a regional medical center contacted individuals who met inclusion criteria; English speaking, > 25 years old and discharged with an MI diagnosis in the last 3-7 years to capture individuals who had transitioned beyond the acute recovery period while remaining at meaningful risk for recurrent events. Excluded were those that had been prescribed antidepressants in the last 3 months, scored above 6 on the Abbreviated Mental Test, had major surgery in the previous 6 months or reported being diagnosed with chronic fatigue syndrome, multiple sclerosis, human immuno-deficiency syndrome, or rheumatoid arthritis. Recruitment efforts included phone contact. Participants were met at a location that was convenient for the participant to avoid transportation as a barrier. Participants completed demographic and health-related questionnaires, including sleep and perceived stress measures. During the visit, a venous blood sample was collected. Blood samples were centrifuged at 1200g and placed in ice until stored in a -20°C freezer.

### Variables and Measures

#### Sleep Quality

Sleep quality was assessed using the 19-item Pittsburgh Sleep Quality Index^48^ (PSQI), which is a self-report measure of various components of sleep quality, duration, latency, disturbances, efficiency, daytime dysfunction and use of sleep medication.^49^ A global total score is also generated. A higher score indicates worse sleep quality with scores above 5 reflecting significant sleep quality.^48^ The PSQI has fair to good internal consistency with Cronbach’s alpha coefficients ranging from .64 to .83.^50^

#### Inflammatory Biomarkers

Inflammatory biomarkers were measured using blood assays. Blood assays were chosen over salivary or urinary analysis due to their superior accuracy in detecting inflammatory markers.^51,52^ Inflammatory markers included in this analysis are IL-1β, IL-6, CRP, and TNFα. Lower levels of detection were calculated, and any concentration values that fell below the lower threshold for detection were coded as 0 pg/ml, rather than missing, as cytokine concentrations were still present.^53^

#### MI recurrence

MI recurrence was measured by the number of MIs participants reported experiencing in the previous 3 to 7 years.

#### Psychological Stress

Psychological stress was assessed using a single Likert-scale investigator-developed question: “During the past month how much stress did you experience in life and work.” Responses ranged from no stress (1) to extreme stress (6). This item was intended to capture a global perception of stress rather than specific item-level stressors. As a single-item measure, it does not permit assessment of internal consistency or differentiation among distinct sources of stress.

#### Covariates

Race, gender, and age were included as covariates due to their established associations with MI risk.^9,10,54^

### Statistical Analyses

An a-priori power analysis was conducted using G*Power v3.1^55^ which indicated that a sample size of *n* = 115 with a power of .95 would yield a significant result at *p* = .05 for a bivariate correlation. This analysis aligns with Fritz et al.^56^ who suggested that a conditional process analysis should have > 100 participants. Further, power issues are uncommon since the analyses involved bootstrapping 1000 times.^57^

Data were analyzed using IBM SPSS v27 (IBM Corporation) and pairwise deletion was completed for missing cases. Descriptive statistics were reported for demographic variables, including age, race, gender, and number of MIs. Correlational analyses were used to describe associations between sleep quality, inflammation, stress, and MI recurrence. In correlational analysis, linear relationships were assumed, and worse sleep quality was expected to be positively associated with higher levels of inflammation. Further, higher numbers of MIs were expected with poorer sleep and higher concentrations of inflammatory biomarkers. After the preliminary analysis, a conditional process analysis^58^ was conducted using PROCESS macro for SPSS (Model 4)^59^ to assess whether sleep quality predicted MI recurrence via different proinflammatory cytokines (IL-1β, IL-6, TNFα, and CRP). We expected higher sleep quality to predict the number of MIs participants experienced, and through conditional pathways involving higher concentrations of inflammatory biomarkers. Finally, a second pathway analyses^58^ (using PROCESS macro for SPSS, Model 8)^59^ tested the interaction effect of psychological stress on the relationship between sleep quality and inflammation predicting MI recurrence. We hypothesized that lower psychological stress would buffer the effects of poor sleep.

## Results

### Demographics

Participants’ (*N* = 156) mean age was 65.37 years (*SD =* 12.13, Range = 34-92) and had experienced an average of 1.48 (*SD =* 0.76) MIs within the last 3 to 7 years. Participants were predominantly White (67.9%, *n =* 106), male (57.1%, *n* = 89), and living with multiple cardiovascular comorbidities (e.g., diabetes, hypertension, hypercholesterolemia; see Table 1). Mean PSQI global scores were 6.58 (*SD* = 3.73; *range* = 2-19) among men and 7.48 (*SD* = 4.00; *range* = 2-16) among women, and 8.11 (*SD* = 3.93; *range* = 2-19) among participants with one MI and 6.32 (*SD* = 3.70; *range* = 2-16) among those with recurrent MIs. Perceived stress scores indicated moderate stress levels in the sample, *M* = 3.10, *SD* = 1.52; range = 1–6.

**Table 1.** Participant Demographic and Clinical Data (N = 156)

|  |  |  |
| --- | --- | --- |
| *Age (years), M= 65.4, SD=12.1* |  |  |
| *Range=34-92* |  |  |
|  | ***n*** | ***%*** |
| *Gender* |  |  |
| Female | 67 | 42.9 |
| Male | 89 | 57.1 |
| *Race/ethnicity* |  |  |
| African American/ | 46 | 29.5 |
| Black | 106 | 67.9 |
| White/Caucasian | 4 | 2.5 |
| Other |  |  |
| *Marital Status* |  |  |
| Married | 98 | 62.8 |
| Single | 16 | 10.3 |
| Divorced/Separated | 27 | 17.3 |
| Widowed | 15 | 9.6 |
| *Highest level of education* |  |  |
| Less than High School | 32 | 20.5 |
| High School Graduate | 47 | 30.1 |
| Technical School | 20 | 12.9 |
| Some College | 29 | 18.6 |
| Bachelor's Degree | 18 | 11.5 |
| Some Graduate School | 3 | 1.9 |
| Graduate Degree | 7 | 4.5 |
| *Medical Problems* |  |  |
| Arthritis | 17 | 10.9 |
| Diabetic | 57 | 36.5 |
| Hypertension | 116 | 74.4 |
| Hypercholesterolemia | 127 | 81.4 |
| Current tobacco use | 36 | 23.1 |
| Peripheral Vascular | 19 | 12.2 |
| Disease |  |  |
| Stroke | 20 | 12.9 |
| *Number of MIs* |  |  |
| 1 MI | 102 | 65.4 |
| 2 or more | 54 | 34.7 |

Note. PSQI = Pittsburgh Sleep Quality Index

### Correlational Analyses

Correlational analyses were conducted to examine bivariate relationships. This analysis provided a preliminary understanding of the direction and strength of relationships between covariate, predictor, and outcome variables, assisted with identifying potential multicollinearity issues between variables, and informed subsequent analysis. Significant associations were observed between sleep quality, inflammation, psychological stress and MI recurrence (see Table 2). Sleep quality was positively correlated with CRP (*r*(143) = .26, *p* = .002), IL-1β (*r*(118) = .19, *p* = .035), and stress (*r*(148) = .30, *p* < .001). Sleep quality was negatively correlated with MI recurrence, (*r*(156) = -.172, *p* = .032). Number of MIs was positively correlated with CRP, (*r*(151) = .24, *p =* .003). Psychological stress was positively correlated with CRP (*r*(150) = .23, *p* = .006), and IL-1β (*r*(125) = .18, *p* = .044). Significant correlations were also detected between cytokines: CRP was positively correlated with IL-6 (*r*(151) = .50, *p* <.001), IL-1β (*r*(126) = .35, *p* < .001), and TNFα (*r*(151) = .23, *p* = .004). IL-6 was positively correlated with TNFα (*r*(151) = .40, *p* < .001). As shown in Table 2, the strongest correlations were observed between sleep and stress, stress and CRP, and between cytokines.

**Table 2.**
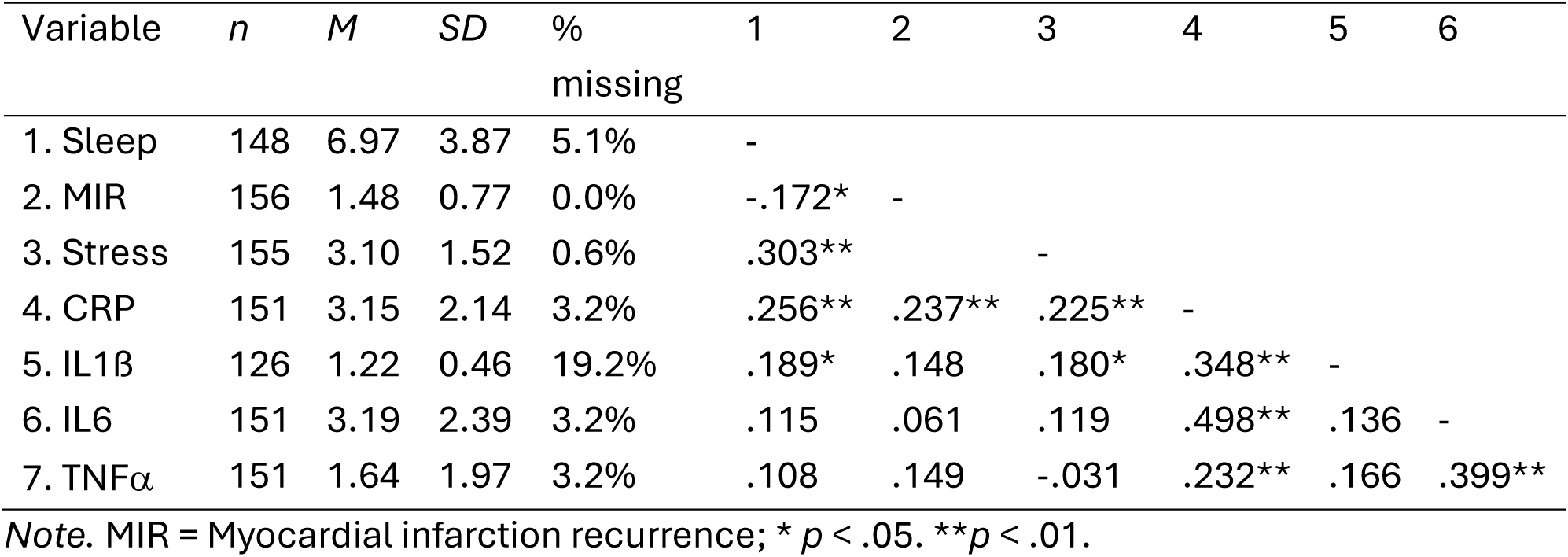
Correlations.

| Variable | *n* | *M* | *SD* | %<br>missing | 1 | 2 | 3 | 4 | 5 | 6 |
| --- | --- | --- | --- | --- | --- | --- | --- | --- | --- | --- |
| 1. Sleep | 148 | 6.97 | 3.87 | 5.1% | - |  |  |  |  |  |
| 2. MIR | 156 | 1.48 | 0.77 | 0.0% | -.172* | - |  |  |  |  |
| 3. Stress | 155 | 3.10 | 1.52 | 0.6% | .303** |  | - |  |  |  |
| 4. CRP | 151 | 3.15 | 2.14 | 3.2% | .256** | .237** | .225** | - |  |  |
| 5. IL1β | 126 | 1.22 | 0.46 | 19.2% | .189* | .148 | .180* | .348** | - | |
| 6. IL6 | 151 | 3.19 | 2.39 | 3.2% | .115 | .061 | .119 | .498** | .136 | - |
| 7. TNFα | 151 | 1.64 | 1.97 | 3.2% | .108 | .149 | -.031 | .232** | .166 | .399** |

Note. MIR = Myocardial infarction recurrence; \* $p < .05$ . \*\* $p < .01$ .

### Sleep Quality and MI Recurrence

A conditional process model (Model 4)^59^ was fit to test the hypotheses that sleep quality directly predicted MI recurrence and indirectly predicted MI recurrence through inflammatory markers. Results indicated that sleep quality did not directly predict MI recurrence, nor did it exert any conditional indirect effects through the inflammatory cytokines examined. However, sleep quality significantly predicted CRP (*b* = 0.16, 𝑟^2^ = 6.93, *p* =.004), explaining approximately 7% of its variance. Sleep quality also predicted IL-1ß (*b* = 0.03, 𝑟^2^= 0.04, *p* =.041) and IL-6 (*b* = 0.11, 𝑟^2^ = .04, *p* =.049), each explaining about 4% of the variance. Despite these relationships, none of the inflammatory cytokines significantly predicted MI recurrence.

### Stress as a Moderator

A conditional process model (Model 8)^59^ was fit to test the hypothesis that stress moderated the pathway linking sleep quality, inflammation, and MI recurrence. The predictor variable, sleep quality, was mean-centered. Results indicated that a combination of the three variables significantly predicted MI recurrence, *F*(3,116) = 8.64, *p* < .005, *R²* = .07. Stress emerged as a significant moderator of the direct effect of sleep quality on MI recurrence. When stress was high, sleep quality predicted MI recurrence (*b* = .07, *p* =.034), whereas this relationship was not significant at low or moderate stress levels. Stress did not moderate the pathway from sleep quality to inflammatory biomarkers. Low, moderate, and high stress correspond to the 16^th^, 50^th^, and 84^th^ percentiles of the stress distribution, which in this sample mapped to 1, 3, and 5, respectively.

## Discussion

The findings from this study provide important insights into the relationship between sleep quality, inflammation, and MI recurrence, with particular emphasis on the role of psychological stress as a moderating factor within the tested pathway. Conditional process analysis provided a robust framework to evaluate both direct and indirect pathways simultaneously while assessing these relationships under specific conditions.

An unexpected finding was the negative bivariate association between sleep quality and MI recurrence, whereby individuals reporting poorer sleep had experienced fewer recurrent myocardial infarctions. This counterintuitive pattern contrasts with established physiological models linking sleep disturbance to adverse cardiovascular outcomes^13,60^ and may reflect the complex clinical context of post-MI populations rather than a protective effect of sleep disturbance. The inverse association observed did not persist in conditional process models, underscoring the context-dependent nature of the relationship and supported cautious, hypothesis-generating interpretations.

Results indicated that sleep quality was significantly correlated with elevated levels of acute and chronic inflammatory markers, including CRP IL-1β, and IL-6, but these inflammatory markers did not explain conditional effects in the relationship between sleep quality and MI recurrence. However, sleep quality was directly associated with increased levels of CRP and IL-1β, suggesting that poor sleep quality may modulate inflammatory cascades, potentially elevating cardiovascular risk and MI recurrence over time. Longitudinal studies are warranted to further elucidate the temporal dynamics of these relationships and explore underlying causal mechanisms.

Psychological stress emerged as a significant moderator within the pathway linking sleep quality to MI recurrence. Psychological stress was assessed using an investigator-developed single-item global appraisal and therefore did not differentiate among specific stressors (e.g., loneliness, grief, illness, or occupational demands) as opposed to a complete, valid, and reliable instrument. Nevertheless, this measure captured individuals’ overall perceived stress burden, which may be a proximal driver of HPA axis activation and downstream inflammatory responses,^61,62^ making it a meaningful indicator for examining stress-related amplification of sleep–cardiovascular pathways. In our sample, high stress levels amplified the impact of poor sleep on MI recurrence, indicating that individuals experiencing both high stress and inadequate sleep may be at particularly elevated risk for subsequent cardiac events.

Furthermore, large global studies, such as Interheart,^54^ have shown that psychological stress can double the risk of cardiovascular events even after adjusting for traditional risk factors. This is especially relevant for aging populations, who often encounter unique stressors such as loneliness, grief and illness.^63,64^ Stress can also precipitate unhealthy behaviors and contribute to cardiovascular risk. Links between stress, dietary patterns, and cardiovascular events through elevated cortisol have been discussed in the literature,^65^ and social stressors such as conflict, loss, or threats have been highlighted as disruptors of sleep and increase inflammation.^66^ While stressors such as loneliness, grief, illness, and caregiving may be particularly relevant in older adulthood, they were not directly measured in our study, nor did we distinguish occupational and non-occupational stressors. Nevertheless, our findings align with this literature, reinforcing the potential value of integrated interventions targeting both sleep quality and stress management to reduce MI recurrence.

It is noteworthy that the expected effect of inflammatory markers in the relationship between sleep and MI recurrence was not observed, suggesting that the pathways linking sleep quality to MI recurrence may be more complex than initially hypothesized and may involve mechanisms beyond the inflammatory markers examined. While sleep loss is known to increase cytokine levels, the cumulative effects of persistent sleep disturbance remain unclear. Irwin^24^ noted that chronic sleep disruption leads to sustained inflammatory responses, which may ultimately contribute to MI recurrence. Chronic inflammation is implicated in major disease processes, including both cardiovascular disease and depression, both relevant to this study. We found that stress moderated the relationship between sleep quality and MI recurrence. Given that older adults may be particularly vulnerable to both stress and impaired sleep, secondary prevention strategies targeting these factors may be especially valuable. Examples include behavioral sleep interventions (e.g., sleep hygiene education or cognitive behavioral therapy for insomnia) and stress-reduction approaches such as mindfulness-based stress reduction or relaxation training, which have demonstrated benefits for sleep quality and psychological well-being in cardiovascular and aging populations.^67–69^

Future research should explore additional biological and psychological pathways that may more fully explain the connection between sleep and cardiovascular outcomes. Prior research has demonstrated associations between poor subjective sleep quality and elevated blood pressure and hypertension, both of which commonly occur in conjunction with MI occurrence.^70,71^ In Irwin’s^24^ proposed pathway linking chronic stress, sleep disturbance, and inflammatory processes, multiple central nervous system mechanisms signal inflammatory responses while modulating immune responses. Our study addresses only a portion of this complex pathway, underscoring the multidimensional nature of MI and its recurrence. Future studies may benefit from incorporating additional covariates and contextual variables such as medication use, comorbidity burden, depressive symptoms, physical activity, and objective sleep characteristics to more fully model these interrelated pathways. Continued research is essential to advance understanding of the interplay between sleep, inflammation, psychological stress, and cardiovascular outcomes.

## Conclusions

This study underscores the complex interplay between sleep quality, inflammation, and psychological stress in predicting MI recurrence. While sleep quality was associated with elevated inflammatory responses, these markers did not account for conditional effects on MI recurrence. Instead, psychological stress emerged as a key moderator, amplifying the impact of poor sleep on cardiovascular outcomes. These findings highlight the importance of comprehensive secondary prevention strategies that address both sleep quality and stress management to reduce the risk of recurrent MIs in aging adults. Further research should prioritize discovering the underlying mechanisms to inform the development of targeted interventions aimed at reducing MI recurrence in this vulnerable population.

## Limitations

There are several limitations to discuss. Data were cross-sectional, retrospective, and collected from a single study site using convenience sampling, which precludes causal inference. The sample was drawn from a specific southeastern region of the U.S. which limits generalizability, particularly in light of regional health inequities and higher mortality rates.^72^ Gender, race, and their intersection are known to influence MI risk. However, this study focused on broader relationships and did not probe multi-level differences in MI recurrence, which may affect statistical significance.

Participants in this study had multiple cardiovascular and metabolic comorbidities, including diabetes, hypertension, and hypercholesterolemia, which are independently associated with systemic inflammation. Although these conditions reflect the clinical reality of post-MI populations, their presence may have influenced baseline cytokine levels and attenuated observed associations between sleep quality, inflammation, and MI recurrence. Future studies should consider incorporating comorbidity indices or disease burden scores to better disentangle the relative contributions of sleep, stress, and underlying medical conditions to inflammatory processes and cardiovascular outcomes.

Although the hypotheses tested in this conditional process analysis are theoretically sound, feedback loops regulating pro-inflammatory cytokines, sleep, psychological stress, and downstream effects such as MI recurrence are complex and often challenging to interpret in regression-based models that specify directional effects. Relationships may also be influenced by cytokines or other variables not included in the model. Self-report bias may have affected measurements of sleep and psychological stress. Additionally, latent constructs such as stress are susceptible to random error, potentially confounding parameter estimates. While the PSQI is a widely used and validated measure of sleep disturbances, it does not provide objective assessments of sleep health or hypersomnolence. Nevertheless, its established validity relative to polysomnography, the gold standard for sleep data, supports its use in evaluating sleep quality in this study.

Another limitation of our study is the use of conditional process analyses rather than traditional mediation analyses, which restricts causal interpretation. While this analytical approach allows simultaneous testing of direct indirect effects, as well as moderating pathways, it remains regression-based and relies on cross-sectional data.^56^ Unlike traditional mediation analyses, which often test indirect effects under fixed assumptions, conditional process models expand this framework by including moderation, but they cannot establish temporal or causal order without longitudinal or experimental data.^57^ Therefore, findings should be interpreted as conditional associations rather than causal mechanisms.

Further research should further explore relationships between biomarkers, sleep, psychological stress, and cardiovascular outcomes such as MI. Studies incorporating theoretically informed covariates, latent variable modeling, and longitudinal or experimental designs will be crucial to clarify these pathways. This study provides a foundational step toward understanding these complex relationships and guiding future investigations.

## Data Availability

All data produced in the present study are available upon reasonable request to the authors

